# Excess cardiovascular deaths in the beginning of COVID-19 outbreak

**DOI:** 10.1101/2020.08.29.20184317

**Authors:** Junren Wang, Jianwei Zhu, Huazhen Yang, Yao Hu, Yajing Sun, Zhiye Ying, Yuanyuan Qu, Unnur A Valdimarsdóttir, Fang Fang, Huan Song

## Abstract

**Importance:** The healthcare demand created by the COVID-19 pandemic was far beyond the hospital surge capacity in many countries, resulting in possible negative influence on prognosis of other severe diseases, such as cardiovascular disease (CVD).

**Objective:** To assess the impact of the COVID-19 outbreak on CVD-related hospitalizations and mortality.

**Design:** Community-based prospective cohort study.

**Setting** the UK Biobank population.

**Participants:** 421,717 UK Biobank participants who were registered in England and alive on December 1^st^ 2019.

**Main outcomes and measures:** The primary outcome of interest was CVD death, as deaths with CVD as a cause of death according to the death registers. We retrieved information on hospitalizations with CVD as the primary diagnosis based on the UK Biobank hospital inpatient data. The study period was between December 1^st^ 2019 and May 30^th^ 2020, and we used the same calendar period of the three preceding years as the reference period. Standardized mortality/incidence ratios (SMRs/SIRs) with 95% confidence intervals were used to estimate the relative risk of CVD outcomes during the study period, compared with the reference period, to control for seasonal variations and aging of the study population.

**Results:** We observed a distinct increase in CVD-related deaths in March and April 2020 as compared to the corresponding months of the three preceding years. The observed number of CVD death (n=217) was almost doubled in April, compared with the expected number (n=120), corresponding to an SMR of 1.81 (95% CI 1.58-2.06). We observed a sharp decline of CVD hospitalization in March (n=841) and April (n=454), compared with the expected number (n=1208 for March and 1026 for April), leading to an SIR of 0.70 (95% CI 0.65-0.74) for March and 0.44 (95% CI 0.40-0.48) for April. There was also a clear increase of death, but a clear decrease of hospitalization, in March and April for all the five major subtypes of CVD.

**Conclusions:** We observed a distinct excess in CVD deaths in the beginning of the COVID-19 outbreak in the UK Biobank population. In addition to CVD complications of SARS-CoV-2 infections, the reduced hospital capacity might have contributed to the observed excess CVD deaths.

**Key Points:** *Question:* How did the COVID-19 outbreak affect rates of CVD-related death and hospitalization?

*Finding:* In this prospective study involving 421,717 UK Biobank participants, we observed excess CVD-related mortality in parallel with decreased CVD-related hospitalization in the beginning of the COVID-19 pandemic, March and April 2020.

*Meaning:* In addition to severe SARS-CoV-2 infection progressing to CVD-related death, reduced hospital resources might have partially contributed to the excess CVD mortality.

## Background

Since the first case of the Corona Virus Disease 19 (COVID-19) in December 2019, the potent contagious disease has spread rapidly all over the globe, with profound influences on the global economy and population health^1^. As of August 2020^2^, with almost 18 million cases and approximately 700,000 deaths, COVID-19 has brought tremendous pressure to the healthcare service systems worldwide. The healthcare demand created by the COVID-19 pandemic is far beyond the hospital surge capacity in many countries, including in the US and the UK^3^. As a result, in addition to insufficient ascertainment and treatment of COVID-19, the severe shortage of medical resources might have considerable impact on patients with other severe diseases. Yet, data illustrating the impact of the COVID-19 outbreak on prognosis of other severe diseases, such as cardiovascular disease (CVD), are limited^4^.

Here, taking advantage of a community-based sample in the UK Biobank with timely and continuously updated longitudinal data on various health outcomes, we estimated the potential impact of the COVID-19 pandemic on CVD-related death and hospitalization.

## Methods

### Study design

Based on the UK Biobank cohort, which has been described in details elsewhere^5^, we included participants (n=421,717) who were registered in England and alive until December 1^st^ 2019, as the inpatient hospital data during the COVID-19 outbreak are currently not available for participants in Scotland and Wales. The study was approved by the biomedical research ethics committee of West China Hospital (reference number: 2020.661).

The primary outcome of our interest was CVD death, which was defined as a death with CVD as a death cause according to the International Classification of Diseases [ICD]-10 codes I00-I70, I730, and I74-I75^6^, based on linked data from the death registers updated until May 30^th^ 2020. In sub-analyses, we focused on five major subtypes of CVD, including acute myocardial infraction, arrhythmia, essential hypertension, heart failure and stroke (ICD codes shown in Supplementary Table 1). The study period was between December 1^st^ 2019 and May 30^th^ 2020. To take into account seasonal variations in CVD mortality^7^, we used the same calendar period of the three preceding years, as the reference period.

Because COVID-19 patients with cardiovascular comorbidity are more likely to experience fatal outcomes^8^ and infection of SARS-CoV-2 leads to cardiac injuries^9,10^, we in a separate analysis excluded individuals with confirmed COVID-19 who died of CVD (i.e., with a positive COVID-19 test according to data from Public Health England, a hospital admission for COVID-19 according to the UK Biobank hospital inpatient data, or a cause of death concerning COVID-19 according to data from the death registers).

As a proxy for access to healthcare for severe CVD during the outbreak, we retrieved information about hospitalizations with inpatient diagnoses of CVD from the UK Biobank hospital inpatient data. Hospitalization for CVD was defined as a hospital admission with CVD as the primary diagnosis during the study period.

We further considered CVD diagnosed before December 1^st^ 2019 as history of CVD, according to the UK Biobank hospital inpatient data.

### Statistical analyses

We first calculated the monthly numbers of CVD death between December 1^st^ 2019 and May 30^th^ 2020. We also calculated the mean monthly numbers of CVD death in the corresponding six months during the three preceding years. To further account for aging, we calculated the expected age-standardized monthly numbers of CVD death for the study period by multiplying the numbers of persons by the average age (1-year strata)-, sex-, and month-specific mortality rates derived from the same period of the three preceding years (i.e., the reference period), based on the same population. The standardized mortality ratios (SMRs), i.e., the ratio of the observed to the expected number of CVD deaths, with its 95% confidence interval^6^ was then used to estimate the relative risk of CVD death, comparing the study period to the reference period. We further performed separate analyses for five major subtypes of CVD, as well as subgroup analyses by history of CVD. We performed the same analyses, including calculations of standardized incidence ratios (SIRs), for CVD hospitalization. All analyses and graph drawing were carried out using R version 3.4.0.

## Results

We observed a sharp increase in the number of CVD death in March and April but not in May 2020, compared with the corresponding months of the three preceding years (Figure 1A and Supplementary Table 2). After taking into account aging of the population during the four years of observation, the observed number of CVD (n=217) was almost doubled, compared with the expected number (n=120), in April (Figure 1B), corresponding to an SMR of 1.81 (95% CI 1.58-2.06) (Figure 1C). After excluding CVD deaths with confirmed COVID-19, we observed a diminished but still statistically significant increase in March (SMR decreased from 1.38 [95% CI 1.17-1.6] to 1.19 [95% CI 1.00-1.40]) but not in April (SMR=0.97, 95% CI 0.80-1.15) (Figure 1C).

**Figure 1.**
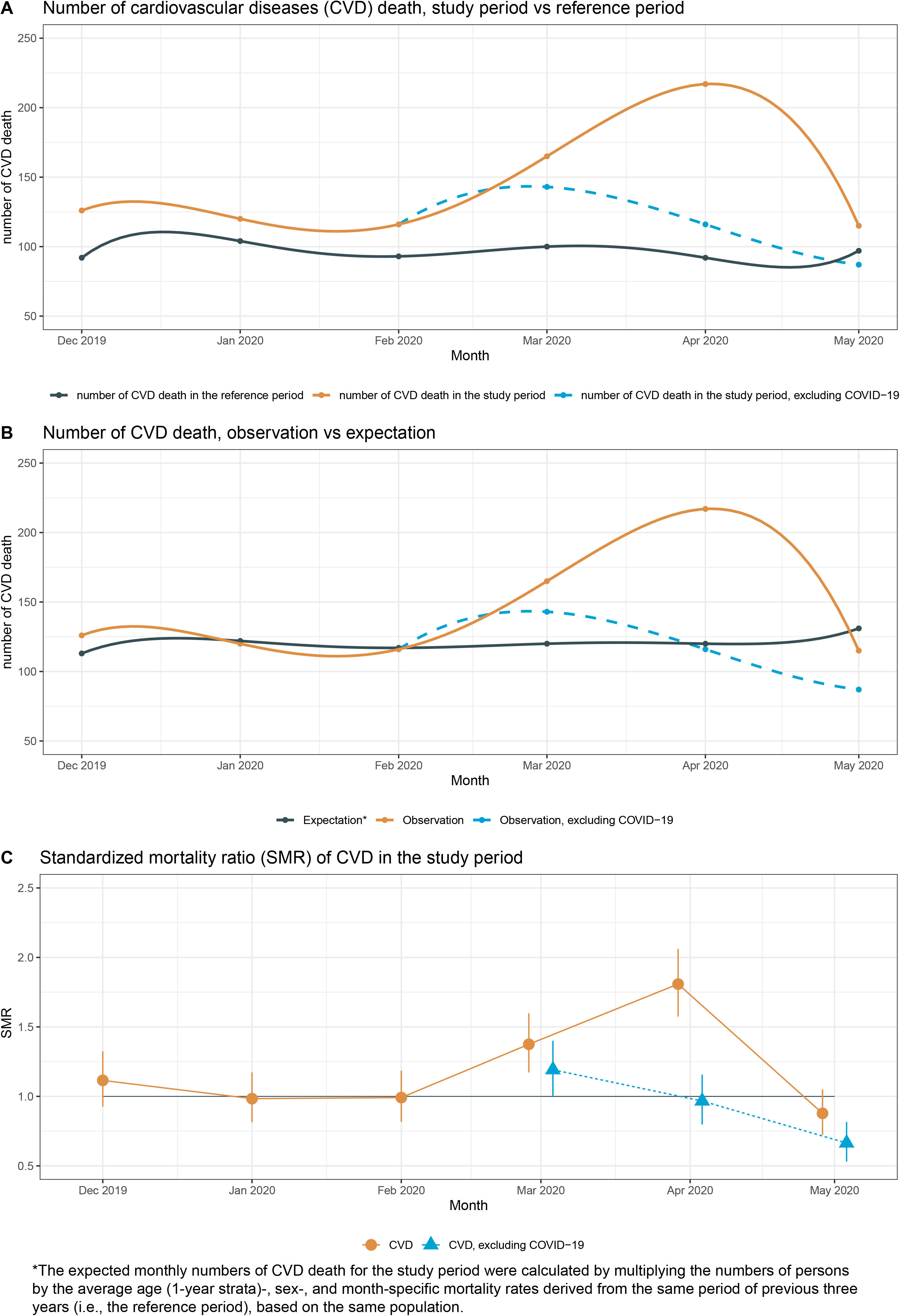
The number and standardized mortality ratio (SMR) with 95% confidence interval of CVD *The expected monthly numbers of CVD death for the study period were calculated by multiplying the numbers of persons by the average age (1 -year strata)-, sex-, and month-specific mortality rates derived from the same period of previous three years (i.e., the reference period), based on the same population.

We observed a rapid decline of CVD hospitalization from March onward, compared with January and February 2020 (Figure 2 and Supplementary Table 3). For both rates of CVD death and hospitalization, the pattern did not differ substantially by history of CVD (Supplementary Figure 1). Figure 3 shows the monthly numbers of CVD death and hospitalization for the five major subtypes of CVD. In general, during the six-month study period, the rates of death peaked for almost all five subtypes of CVD in March or April (Figure 3A). The peak in CVD mortality diminished to some extent, but remained evident, after excluding CVD deaths with confirmed COVID-19 (Figure 3B). In contrast, the rates of CVD hospitalization were lowest during March or April for all five subtypes of CVD (Figure 3C).

**Figure 2.**
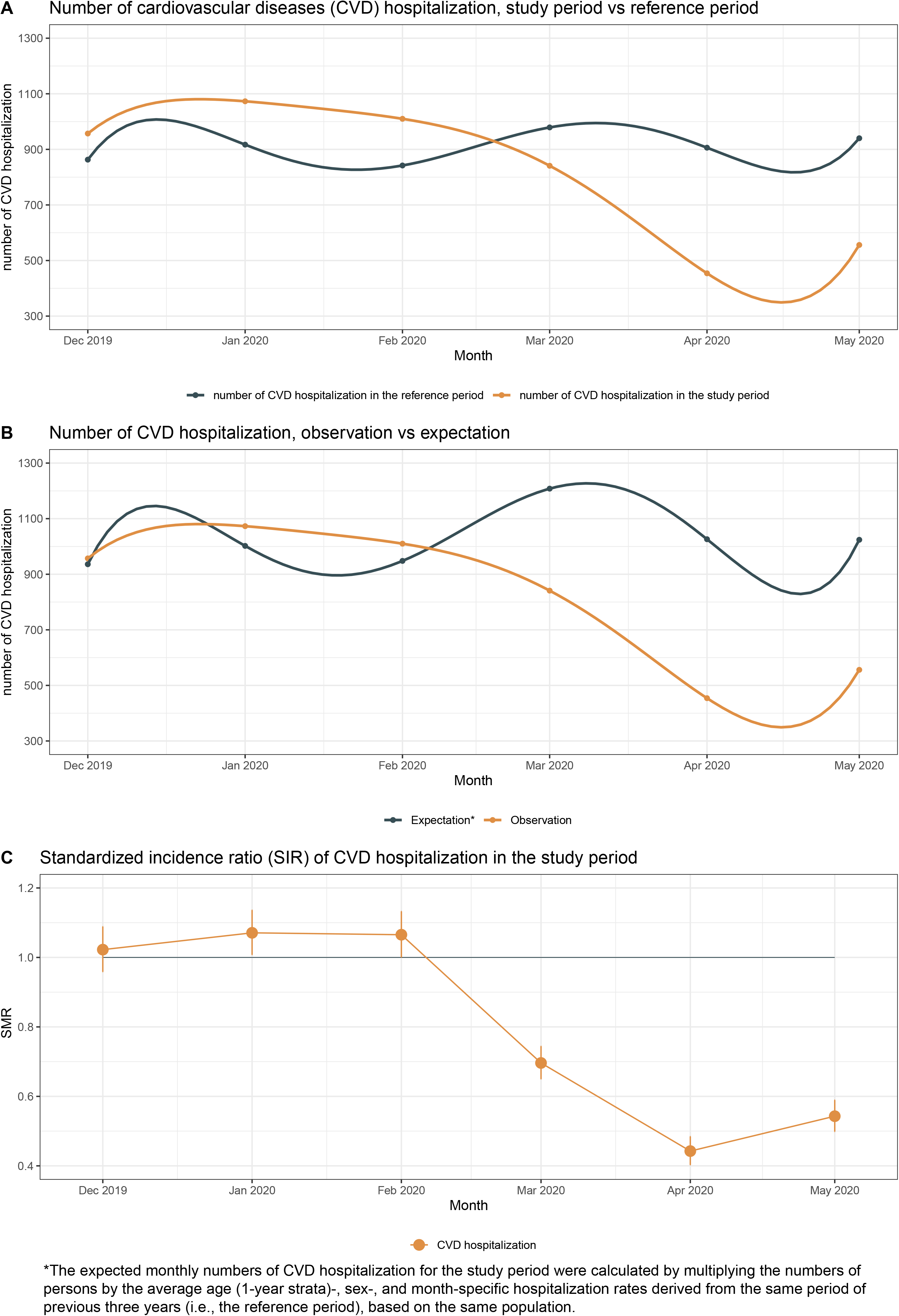
The number and standardized incidence ratio (SIR) with 95% confidence interval of CVD hospitalization *The expected monthly numbers of CVD hospitalization for the study period were calculated by multiplying the numbers of persons by the average age (1-year strata)-, sex-, and month-specific hospitalization rates derived from the same period of previous three years (i.e., the reference period), based on the same population.

**Figure 3.**
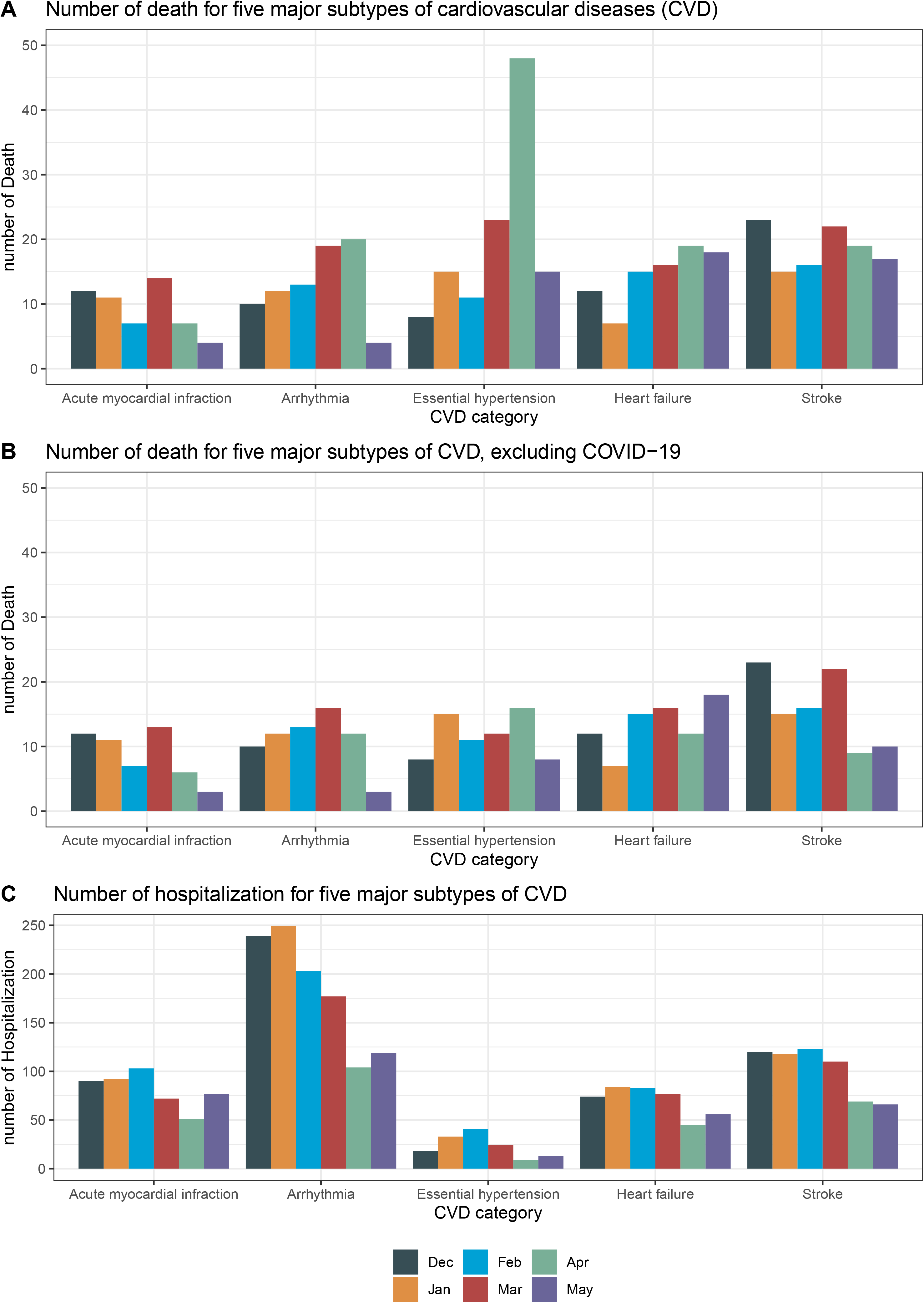
The number of CVD death and hospitalization for five major subtypes of CVD

## Discussion

Based on analyses of a large community-based cohort study using the UK Biobank data, we found a considerably increased risk of CVD-related deaths, while in parallel a significantly reduced rates of CVD hospitalization, in March and April 2020. This matches the exponential growth of COVID-19 cases in the UK during the time (e.g., from 35 to 25,521 from beginning to the end of March), suggesting that the shortage of healthcare resources for regular CVD care might have partially contributed to the excess CVD mortality. Similarly, the fear of accessing health care by patients in need might have led to delayed treatment, contributing further to an increase in CVD death. Severe SARS-CoV-2 infection has been associated with cardiac injuries^11^ which may also contribute to the increased CVD-related mortality. However, after exclusion of CVD deaths with a known COVID-19, the excess CVD mortality remained statistically significant in March, especially among participants without history of CVD. The null result in April might reflect the greater spread of COVID-19 affecting individuals more prone to fatal cardiovascular events in April than March^12^. Further, the greater risk of CVD death in March and April, compared with May when the COVID-19 was still largely spreading in the UK, implies that other factors, such as severe emotional stress reactions to the COVID-19 outbreak which have been demonstrated as a trigger for CVD incidence and mortality in relation to other stressful life events^6^, may also contribute to the observed excess deaths of CVD.

The major merit of our study is the self-comparison analysis, contrasting the numbers and rates of CVD death and hospitalization during the study period (COVID-19 outbreak), with the corresponding numbers and rates of a reference period (the same calendar period during the three preceding years), using the UK Biobank population as the same study population. This analysis adjusted inherently for potential confounders that are constant over time, including genetic factors and many environmental factors that did not change greatly within the 4-year period. Other influential factors, such as seasonal variation and aging of the study population, were well controlled by monthly-based estimations and calculation of standardized expected numbers and SIRs.

The notable limitations of this study include the fact that accessibility to COVID-19 test in the UK was largely restricted to inpatients with symptoms before May 2020. Consequently, we might have under-estimated number of deaths related to COVID-19 in March and April. In addition, the UK Biobank population is not representative of the general population in the UK. It therefore requires cautions when generalizing our findings to the whole UK population or other populations.

## Conclusions

Excess deaths from CVD were observed in the UK Biobank population in March and April 2020. CVD complications of COVID-19 but also decreased capability of hospital healthcare system might have contributed to the observed excess risk of CVD death.

## Data Availability

Data sharing: Data from UK Biobank (http://www.ukbiobank.ac.uk/) are available to all researchers upon application. This research has been conducted using the UK Biobank Resource under Application 54803.

## Acknowledgements

We thank the team members involved in West China Biomedical Big Data Center for Disease Control and Prevention for their support.

## Funding

This work is supported by the National Science Foundation of China (No. 81971262 to HS), West China Hospital COVID-19 Epidemic Science and Technology Project (No. HX-2019-nCoV-014 to HS), and Sichuan University Emergency Grant (No. 2020scunCoVyingji1002 to HS).

## Role of Funder/Sponsor

The sponsors had no role in the design and conduct of the study; management, analysis, and interpretation of the data; preparation, review, or approval of the manuscript; and decision to submit the manuscript for publication.

## Author contributions

HS, JZ, FF, and UAV were responsible for the study concept and design. YH, HY, ZY, JS, and YQ did the data and project management. JW and HS did the data analysis. JW, HS, FF, JZ, and UAV interpreted the data. JW, HS, FF, JZ, HY, YH, YS, ZY, YQ and UAV drafted the manuscript.

## Competing interests

Authors declare no competing interests.

## Data sharing

Data from UK Biobank (http://www.ukbiobank.ac.uk/) are available to all researchers upon application. This research has been conducted using the UK Biobank Resource under Application 54803.

## Notes

### Competing Interest Statement

The authors have declared no competing interest.

### Author Declarations

The study was approved by the biomedical research ethics committee of West China Hospital (reference number: 2020.661).

